# Wastewater Treatment Plants as Representative Sentinel Sites in Infectious Disease Surveillance

**DOI:** 10.64898/2026.08.27.26361522

**Authors:** Edem Fiatsonu, Dustin Hill, Christopher Dunham, David A. Larsen

## Abstract

Wastewater-based epidemiology (WBE) has emerged as a powerful population-level surveillance tool, but its coverage is structurally concentrated in in-network urban areas, potentially leaving rural populations underrepresented. Routine human movement between sewered (in-network) and unsewered (off-network) areas may, however, cause wastewater treatment plant (WWTP) measurements to reflect infectious disease dynamics beyond sewer boundaries. We evaluated this hypothesis using daily clinical COVID-19 testing data (January 2021–April 2022) across New York State excluding New York City (NYC). We disaggregated weekly cases and tests into in-network (WWTP catchment area) and off-network (outside WWTP catchment area) components applied to two geographic frameworks: administrative counties (N = 53 mixed-coverage) and mobility-defined communities identified through Walktrap community detection applied to census tract-level movement networks (N = 32 mixed-coverage). In/off-network COVID-19 trends were strongly correlated under both frameworks. County-level statewide aggregate correlations were high (incidence r = 0.994, positivity r = 0.996), as were individual county correlations (median r = 0.909 and 0.932, respectively). Mobility-defined community-level statewide correlations were similarly strong (r = 0.990 and 0.992), with comparable unit-level medians (r = 0.877 and 0.894). The mobility-defined community framework provided better population balance between in-network and off-network strata (87.5% vs. 69.8% in balanced range) and a higher floor on representativeness (minimum r = 0.440 vs. 0.177). Population size was the dominant predictor of in-network/off-network alignment at both scales; wastewater infrastructure density and off-network signal variability provided additional explanatory power at the mobility-defined community level. WWTPs broadly represent COVID-19 dynamics in surrounding off-network populations, supporting their use as sentinel surveillance sites. Representativeness weakens in smaller, more rural communities, and mobility-defined communities provide a complementary framework for identifying where this occurs.

**Significance Statement:** Municipal wastewater samples can be tested for infectious diseases circulating in sewered populations, but most surveillance systems exclude rural communities lacking sewer connections. Using clinical COVID-19 data from New York State, we demonstrate that routine human movement between in-network and off-network areas causes wastewater signals to reflect disease dynamics across sewer boundaries. In-network and off-network epidemic trends were highly correlated across both counties and mobility-defined communities, and testing wastewater can effectively surveil infectious diseases among populations beyond the limits of wastewater infrastructure. However, representativeness weakens in smaller communities, and 42% of mobility-defined communities have no wastewater infrastructure. Expanding wastewater surveillance to rural facilities should be treated as a surveillance-equity priority to ensure equitable early-warning capabilities for all communities.

## Introduction

Wastewater-based epidemiology (WBE) is historically useful for poliomyelitis (1) and then expanded as a population-level infectious disease surveillance approach during the COVID- 19 pandemic (2, 3). WBE has been recently applied to respiratory syncytial virus (RSV) (4), influenza A (5), and other pathogens in addition to SARS-CoV-2 as multiple infectious disease pathogens can be detected in wastewater (6). Measuring the amount of a pathogen’s RNA in wastewater enables understanding of a pathogen’s transmission dynamics in the mobility community.

The associations between wastewater pathogen concentrations and clinical indicators have been demonstrated across multiple pathogens where studies have shown strong correlations with reported cases, hospitalizations, and test positivity at sewershed and municipal scales (7–10). In many settings, changes in wastewater viral concentrations have been observed to precede trends identified through clinical surveillance, highlighting the potential of WBE as an early warning tool for mobility community transmission of infectious diseases (9, 10).

Despite these successes, an important unresolved question remains: to what extent do wastewater samples taken from wastewater treatment plants (WWTPs) represent infectious disease dynamics in nearby off-network communities? This question is particularly salient in populations with mixed sanitation infrastructure, where urban populations are largely connected to centralized sewer systems while surrounding rural or peri-urban populations rely on decentralized systems such as septic tanks. In such contexts, concerns have been raised about the equity and completeness of wastewater in public-health surveillance (11).

Rural–urban health disparities are well documented across diverse settings. Rural populations often experience higher disease burdens, reduced access to healthcare, and poorer health outcomes compared with urban populations, driven by structural differences in health system design, socioeconomic conditions, and infrastructure availability (12–15). The lack of a wastewater treatment plant that can provide information for wastewater surveillance in rural areas is potentially one more health disparity that these communities face.

Although rural wastewater does not flow to urban WWTPs, it is plausible that urban wastewater reflects transmission dynamics of both urban and rural communities that are connected through human movement patterns. Individuals routinely travel across sewershed boundaries for employment, healthcare, education, entertainment, and social activities, creating pathways for disease transmission independent of sewer infrastructure. Through human movement patterns, infections acquired in off-network areas may ultimately contribute to wastewater signals measured in sewered communities, suggesting that a WWTP’s data may indirectly reflect infectious disease dynamics beyond the sewer network (11).

Mechanistic modeling studies provide further evidence that pathogens move independently of sewer infrastructure. Using an agent-based model approach integrating SEIR transmission dynamics, GIS-resolved sewer infrastructure, and cross-zone mobility, Xiang et al. demonstrated that in the context of high connectivity, an outbreak originating in non-sewered zones would be detected by WWTP surveillance approximately 1–2 days later than an outbreak originating in sewered areas (17). Importantly, mobility only influenced detection timing when outbreaks began in non-sewered areas, indicating that human movement governs when outbreaks originating in off-network communities become visible in wastewater. Other agent-based and behavioral modeling studies similarly indicate that mobility and mixing between communities influence where and when viral signals emerge within sewer networks (18,19). Although these modeling studies clarify the mechanisms through which mobility and infrastructure influence wastewater surveillance, empirical evaluations that integrate real-world human movement data with wastewater and clinical surveillance remain limited. Large-scale evidence quantifying how mobility-defined communities spanning sewered and off-network areas affect epidemiological alignment remains limited.

In this study, we evaluate whether WWTP measurements reflect infectious disease dynamics in nearby off-network communities through an integrated analysis of wastewater surveillance, clinical epidemiology, and human movement networks. We quantify epidemiological alignment between sewered and off-network areas using clinical incidence, testing metrics, and wastewater signals. Although we use SARS-CoV-2 as a data-rich test case, the question of whether WWTPs represent off-network populations applies broadly to any respiratory pathogen monitored through wastewater. By grounding representativeness in observed mobility rather than infrastructure alone, this work advances WBE toward more equitable and informative understanding of surveillance across rural–urban divides.

## Results

Over the study period (December 27, 2020, to April 24, 2022), 33.0 million COVID-19 tests were conducted across 57 New York State counties outside NYC, yielding 1.98 million positive cases (overall positivity = 6.0%). Of these, 15.2 million tests (46.1%) and 857,187 cases (43.3%) were among individuals residing within catchment areas of WWTP permitted to discharge at least one million gallons per day (in-network), while 17.8 million tests (53.9%) and 1.12 million cases (56.7%) were among the off-network population. The study period captured four distinct epidemic phases: the post-holiday winter surge of 2020–2021, a period of lower transmission through mid-2021, the Delta wave beginning in late summer 2021, and the Omicron surge in winter 2021–2022, which produced the highest weekly incidence and positivity rates observed during the study period.

### County-Level Alignment of COVID-19 Indicators Among On- and Off-Network Communities

Among 57 New York State counties, 54 were mixed-coverage, 2 were entirely off-network (no wastewater treatment plant with at least 1.0 million gallons per day discharge capacity), and 1 fully in-network. At the statewide aggregate level, in/off-network weekly incidence and test positivity tracked closely across all epidemic phases, with very strong correlations (r = 0.994 and r = 0.996, respectively; p < 0.001 for both; Figure 1A). Cross-correlation analysis confirmed synchronous dynamics with peak correlation at lag 0 for both metrics (Figure 1B). At the individual county level, in/off-network incidence correlations were high (median r = 0.909, range: 0.177–0.992; 84.9% exceeding r = 0.80), as were test positivity correlations (median r = 0.932, range: 0.703–0.996; 94.2% exceeding r = 0.80). Weighted regression models identified log-transformed population as the dominant predictor of county-level representativeness (Table 2). Log(population) was significant in all models (incidence: β = 0.427, p < 0.001, adjusted = 0.506; positivity: β= 0.395, p < 0.001, adjusted = 0.654), with the balance index borderline significant for positivity (β= 0.628, p = 0.058). No other predictors reached significance at the county level.

**Figure 1.**
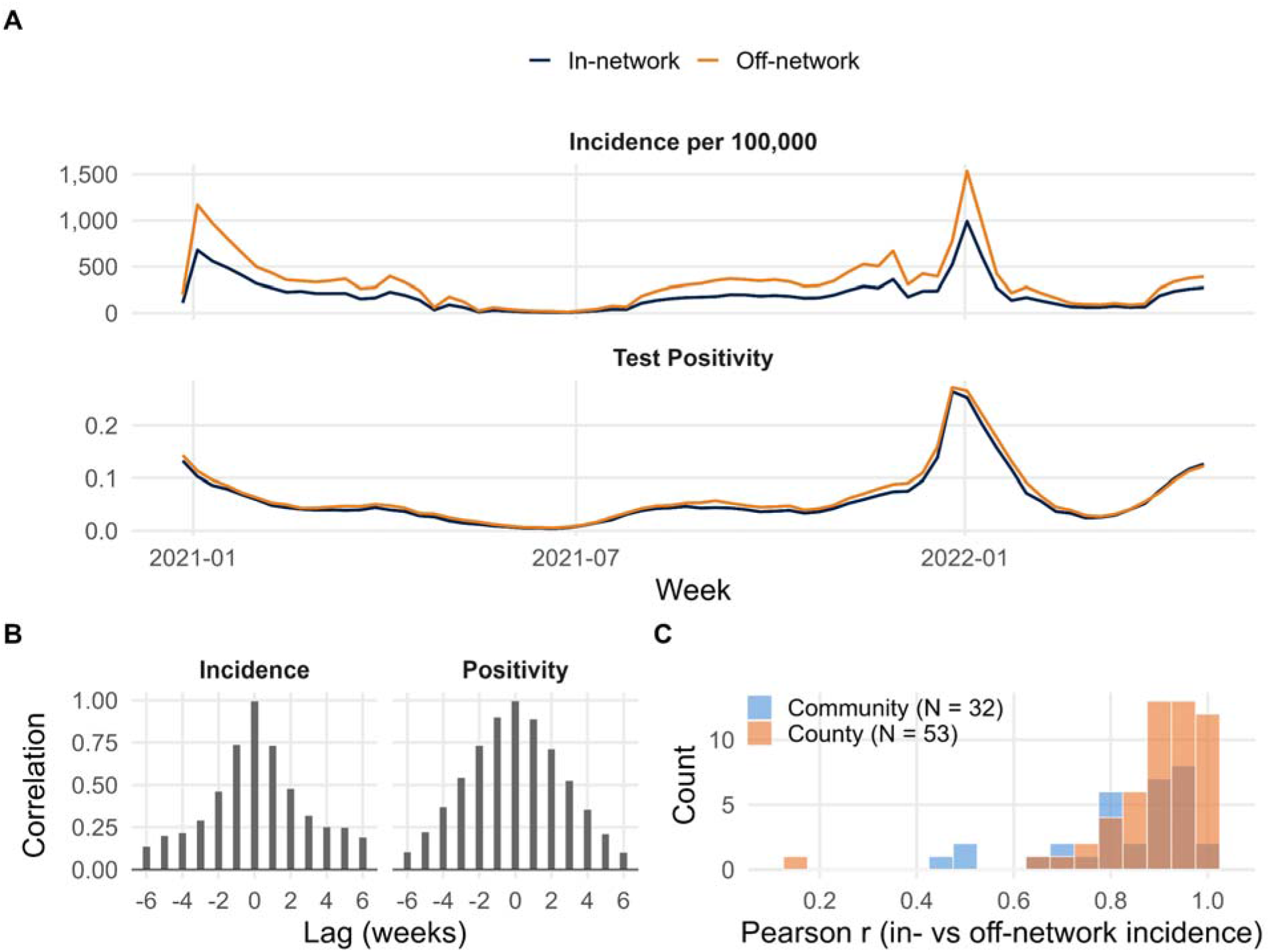
Statewide temporal alignment between in/off-network COVID-19 burden. (A) Weekly in-network (blue) and off-network (orange) COVID-19 incidence per 100,000 and test positivity at the statewide aggregate level across all epidemic phases, January 2021–April 2022. (B) Cross-correlation function for statewide aggregated in/off-network incidence and test positivity at lags ±6 weeks; peak correlation at lag 0 indicates synchronous dynamics. (C) Distribution of unit-level Pearson correlations between weekly in/off-network incidence for mixed-coverage counties (N = 53, orange) and mobility-defined communities (N = 32, blue).

### Mobility-defined community results

Following the exclusion of two Seneca Nation boundary tracts documented in the methods, the Walktrap algorithm produced 64 mobility-defined communities across New York State, encompassing a total population of 10.6 million residents. Of 2,870 census tracts in the shapefile, 2,413 (84.1% of census tracts, 95.4% of population) were successfully assigned to a mobility community based on network connectivity; the remaining 457 tracts (15.9% of census tracts, 4.6% of population) had insufficient mobility connections to be assigned and were excluded from mobility-defined community-level analyses. Unassigned tracts were concentrated in low-density, sparsely populated tracts with limited commercial activity. Among assigned tracts, 2,242 (98.0%) had complete mobility data across all 16 study months; the remaining 45 tracts (2.0%) had incomplete temporal coverage but were retained in community assignments as their limited connectivity contributed minimal weight to the Walktrap algorithm’s community detection. Communities varied substantially in total population and were highly over-dispersed, ranging from 1,148 to 1,939,151 residents (median = 16,413), reflecting New York State’s diverse settlement geography from sparsely populated rural areas to dense urban centers. The sewered fraction defined as the proportion of community population served by WWTPs ranged from 0 to 1 (median = 0.247), indicating that the typical mobility-defined community had approximately one quarter of its population connected to WWTPs participating in the state’s wastewater surveillance network. Based on sewered fraction, communities were classified into three types: 1) 27 communities (42% of mobility-defined communities) representing 130,861 residents (1.2% of the population) were entirely off-network; 2) 5 communities (8%) representing 3,806,111 residents (35.8%) were entirely in-network; and 32 communities (50%) representing 6,680,855 residents (62.9%) were mixed-coverage.

Figure 2 shows the spatial relationship between mobility-defined communities, county boundaries, and sewershed catchments. Mobility-defined community boundaries frequently crossed county lines (Figure 2A-B), indicating cross-jurisdictional connectivity, while sewershed catchments remained largely within county borders. Within mixed-coverage communities (Figure 2B), in-network urban cores and surrounding off-network tracts belonged to the same mobility-defined community, reflecting routine movement between town centers and rural peripheries. Predominantly rural communities, such as those in Essex County (Figure 2C), lacked any connection to in-network sewersheds.

**Figure 2.**
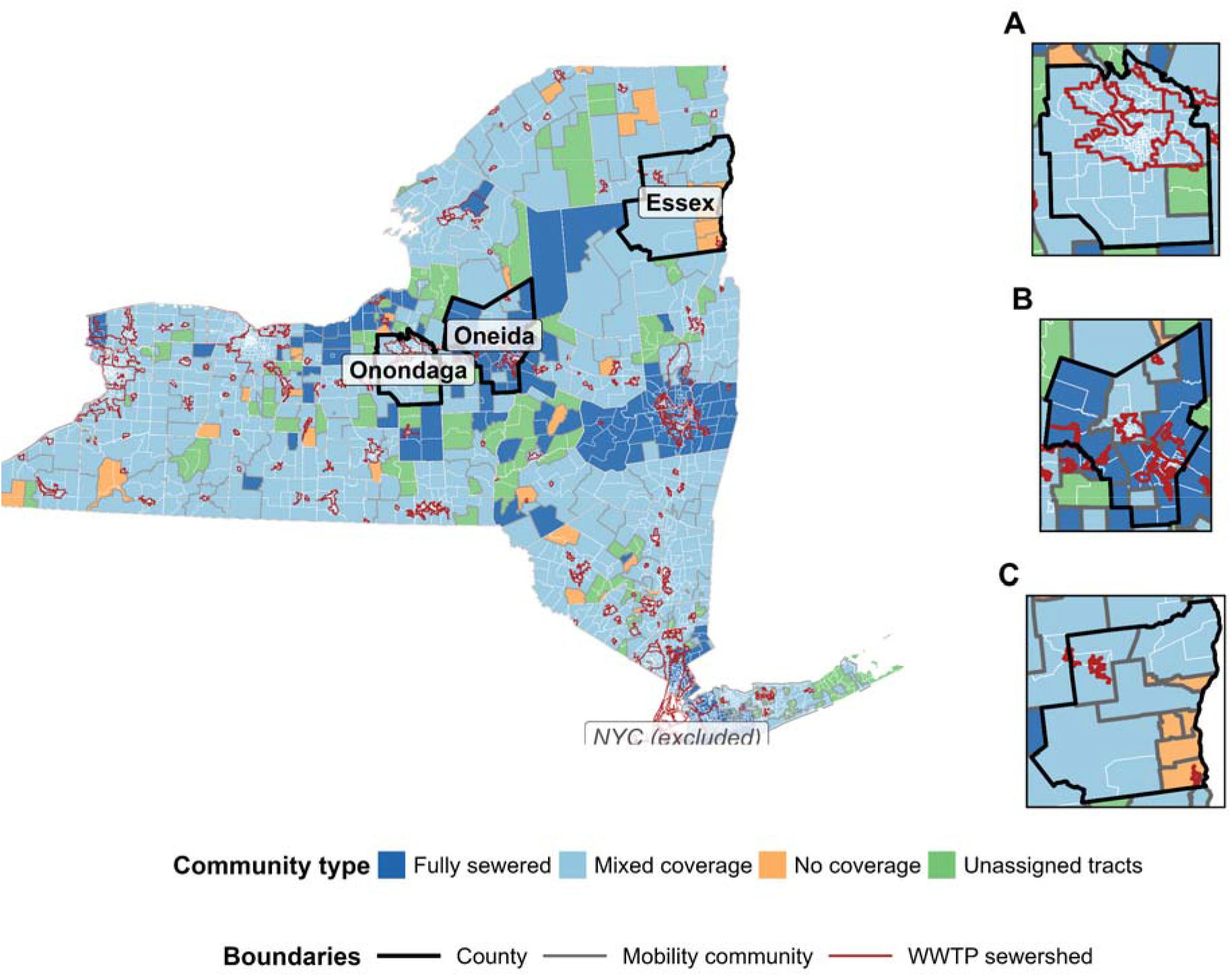
Mobility-defined communities, county boundaries, and WWTP sewershed catchments across NYS outside NYC. The statewide overview shows census tracts colored by mobility-defined community type: fully in-network (dark blue), mixed coverage (light blue), no coverage (orange), and unassigned tracts with insufficient mobility data (green). NYC boroughs, excluded from all analyses, are shown in gray. Outlined counties are shown in zoomed detail in panels *A*–*C*. (*A*) Onondaga County: an urban setting where WWTP sewershed boundaries (red) cross county lines (black), while mobility community boundaries (gray) capture this cross-jurisdictional connectivity. (*B*) Oneida County: a mixed-coverage mobility-defined community with an in-network urban core surrounded by off-network rural tracts, all linked within a single mobility-defined community. (*C*) Essex County: a predominantly rural area with minimal sewershed coverage, exemplifying the 42% of identified mobility-defined communities with no WWTPs.

### Temporal Stability of Mobility-defined communities

To assess the temporal robustness of the detected mobility-defined communities, we computed month-to-month partition similarity across the 16-month study period using two complementary metrics: the adjusted Rand index (ARI) and normalized mutual information (NMI). Both metrics remained consistently high throughout the study period, with ARI ranging from approximately 0.64 to 0.86 and NMI ranging from approximately 0.79 to 0.93 (Figure 3A), indicating that the mobility-defined community structure detected by the Walktrap algorithm was stable across the study period. A transient dip in both metrics was observed in July–August 2021, potentially reflecting shifts in population mobility patterns during the emergence of the Delta variant, when behavioral responses to a novel COVID-19 variant wave may have temporarily altered routine movement patterns. The consistently high ARI and NMI values across the full study period confirm that the mobility-based clustering approach produced coherent and stable population structures, supporting their use as meaningful and reproducible analytical units for epidemiological surveillance.

**Figure 3.**
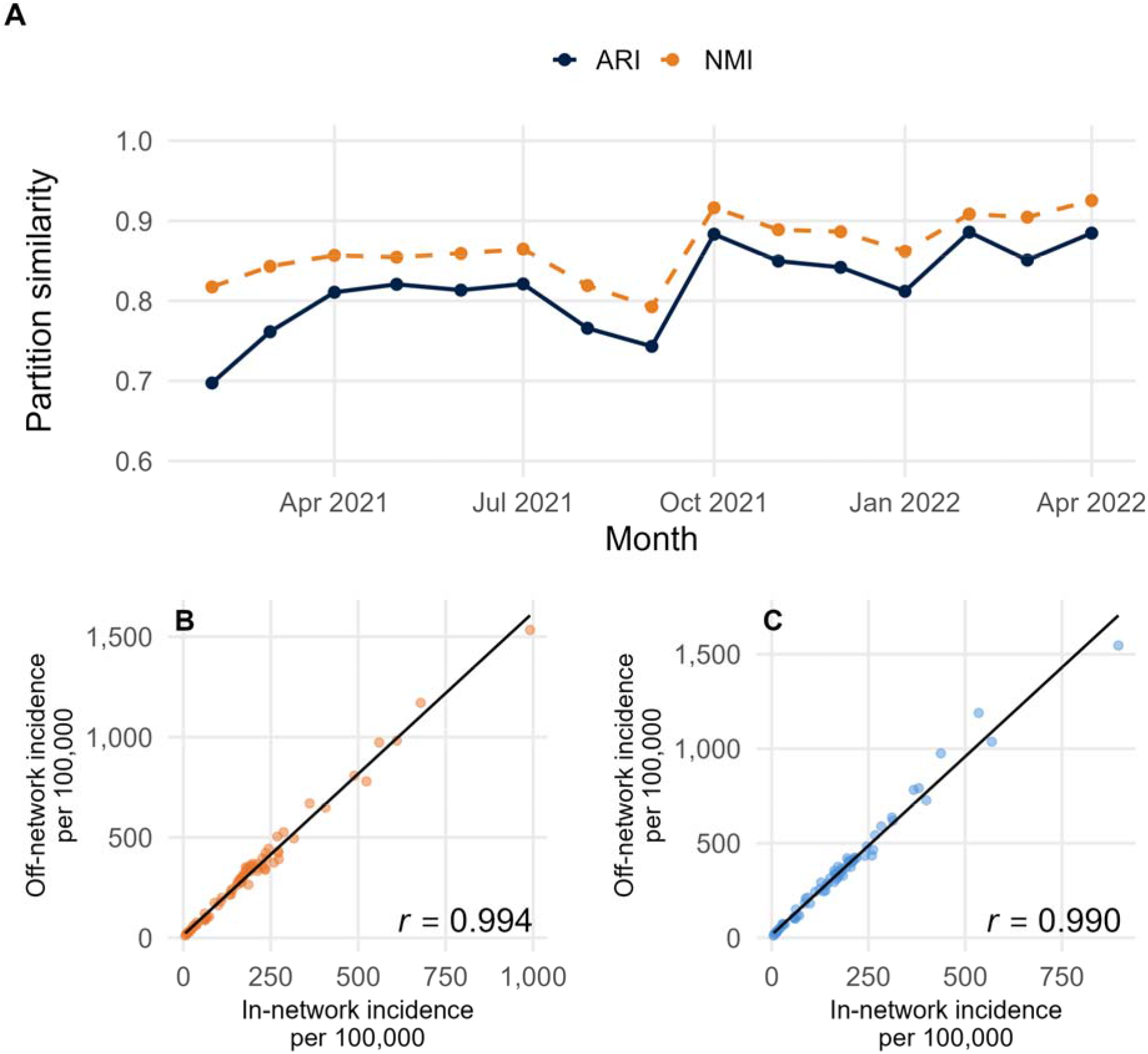
Temporal stability of mobility-defined communities and epidemiological alignment. (*A*) Month-to-month partition similarity across the 16-month study period expressed as adjusted Rand index (ARI; solid line) and normalized mutual information (NMI; dashed line). (*B*) Scatter plot of weekly in-network versus off-network COVID-19 incidence per 100,000 for mixed- coverage counties (r = 0.994). (*C*) Corresponding scatter plot for mixed-coverage mobility- defined communities (r = 0.99).

### Epidemiological Alignment Between In-Network and Off-Network Populations

At the statewide aggregate level, mobility-defined community in/off-network incidence and test positivity were strongly correlated (r = 0.990 and r = 0.992, respectively; p < 0.001; Figure 1A), comparable to the county-based correlations (Fisher z-test: incidence 1.48, 0.138; positivity 2.01, 0.044). The difference in test positivity is attributable to the mobility-defined community framework’s more asymmetric population split (7.66 million in-network versus 2.96 million off-network, compared to the county framework’s near-equal split of 5.68 million versus 5.46 million), producing somewhat noisier off-network rate estimates under the mobility-derived framework. Cross-correlation analysis confirmed synchronous dynamics at lag 0 under both frameworks (Figure 1B). Scatter plots of weekly in-network versus off-network incidence showed tight linear relationships at both scales (county r = 0.994, mobility-defined community r = 0.990; Figure 3B–C). At the mobility-defined community, correlations varied substantially across units, with smaller communities exhibiting weaker and more variable alignment. The median mobility-defined community incidence correlation was 0.877 (range: 0.440–0.990), and the median positivity correlation was 0.894 (range: 0.703–0.996).

### Comparison of county and mobility-defined community frameworks

To evaluate whether mobility-defined communities provided a more appropriate analytical unit, we compared the two frameworks on population balance and incidence alignment (Table 1). While county-level unit correlations were somewhat higher than mobility-defined community correlations (median r = 0.909 versus 0.877 for incidence; Wilcoxon p = 0.040), this difference was modest in absolute terms (bootstrap median difference = 0.035, 95% CI: −0.010 to 0.113). Mobility-defined communities exhibited a higher median balance index (0.363 versus 0.282), with 87.5% falling within the balanced range of 20–80% in-network coverage compared to 69.8% of counties. Notably, the mobility-defined community framework produced a substantially higher floor on representativeness (minimum r = 0.440 versus 0.177), indicating that no mobility-defined community was as poorly served by wastewater surveillance as the worst-performing counties. While nearly all counties (95%) contained at least one WWTP participating in the state network, a large share of mobility-defined communities had no WWTP coverage at all, revealing a rural surveillance gap not apparent when analyzed at the county level.

**Table 1:** County versus mobility-defined community frameworks: incidence alignment

|  | Median sewered fraction | Median balance index | % in balanced range (0.2–0.8) | Median r (incidence) | Min r | SD r | % with r > 0.80 |
| --- | --- | --- | --- | --- | --- | --- | --- |
| County | 0.344 | 0.282 | 69.8 | 0.909 | 0.177 | 0.130 | 84.9 % |
| Mobility-defined community | 0.432 | 0.363 | 87.5 | 0.877 | 0.440 | 0.144 | 71.9% |

### Predictors of Epidemiological Alignment

To identify factors associated with the strength of epidemiological alignment, we fit identical weighted linear regression models at both the county (Table 2) and mobility-defined community (Table 3) levels using Fisher z-transformed correlation coefficients as outcomes, separately for incidence and test positivity. At the county level, log-transformed population was the dominant predictor in both models (incidence: β = 0.444, p < 0.001; positivity: β = 0.371, p < 0.001), with the balance index also significant for test positivity (β = 0.649, p = 0.038; Table 2). At the mobility-defined community level, log-transformed population was again the dominant predictor of incidence alignment (β = 0.505, p < 0.001; Table 3). The test positivity model identified two additional significant predictors: WWTP density (β = 0.089, p = 0.020) and variability in off-network test positivity (β = −8.969, p = 0.012), both retained in the reduced model. While population size dominated at both scales, secondary predictors were scale-dependent: balance index was significant only at the county level, while WWTP density and off-network signal variability were significant only at the mobility-defined community, suggesting that infrastructure characteristics are actionable determinants of representativeness detectable only when geographic units align with population movement patterns.

**Table 2.**
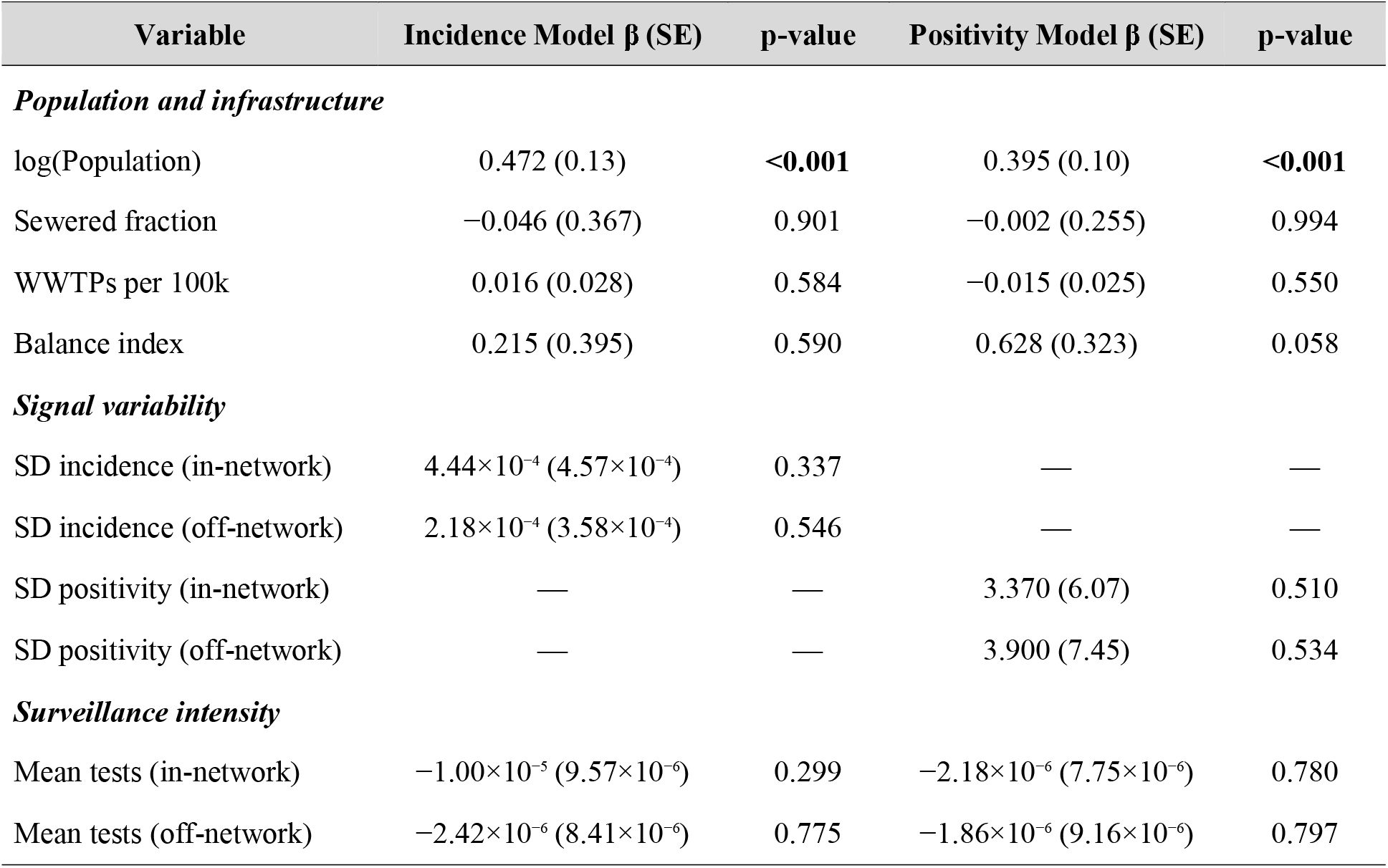
County-level weighted linear regression models predicting Fisher z-transformed in/off-network COVID-19 correlation, NYS. Models restricted to 54 mixed-coverage counties (sewered fraction >0.001 and <0.999). Weights equal the number of complete paired observation weeks per county (minimum = 1). Incidence model: R² = 0.643, adjusted R² = 0.506, residual SE = 2.847 (df = 44), F = 7.66 (8, 44), p < 0.001. Positivity model: R² = 0.728, adjusted R² = 0.654, residual SE = 2.310 (df = 44), F = 13.05 (8, 43), p < 0.001. Bold p-values indicate statistical significance (p < 0.05). WWTP, wastewater treatment plant; SD, standard deviation.

| Variable | Incidence Model $\beta$ (SE) | p-value | Positivity Model $\beta$ (SE) | p-value |
| --- | --- | --- | --- | --- |
| **Population and infrastructure** |  |  |  |  |
| log(Population) | 0.472 (0.13) | **<0.001** | 0.395 (0.10) | **<0.001** |
| Sewered fraction | -0.046 (0.367) | 0.901 | -0.002 (0.255) | 0.994 |
| WWTPs per 100k | 0.016 (0.028) | 0.584 | -0.015 (0.025) | 0.550 |
| Balance index | 0.215 (0.395) | 0.590 | 0.628 (0.323) | 0.058 |
| **Signal variability** |  |  |  |  |
| SD incidence (in-network) | 4.44×10<sup>-4</sup> (4.57×10<sup>-4</sup>) | 0.337 | — | — |
| SD incidence (off-network) | 2.18×10<sup>-4</sup> (3.58×10<sup>-4</sup>) | 0.546 | — | — |
| SD positivity (in-network) | — | — | 3.370 (6.07) | 0.510 |
| SD positivity (off-network) | — | — | 3.900 (7.45) | 0.534 |
| **Surveillance intensity** |  |  |  |  |
| Mean tests (in-network) | -1.00×10<sup>-5</sup> (9.57×10<sup>-6</sup>) | 0.299 | -2.18×10<sup>-6</sup> (7.75×10<sup>-6</sup>) | 0.780 |
| Mean tests (off-network) | -2.42×10<sup>-6</sup> (8.41×10<sup>-6</sup>) | 0.775 | -1.86×10<sup>-6</sup> (9.16×10<sup>-6</sup>) | 0.797 |

**Table 3.** Mobility-defined community-level weighted linear regression models predicting Fisher z-transformed in/off-network COVID-19 correlation, NYS. Models restricted to 32 mixed-coverage communities (sewered fraction >0.001 and <0.999). Weights equal the number of complete paired observation weeks per community (minimum = 1). Incidence model: R² = 0.641, adjusted R² = 0.516, residual SE = 3.093 (df = 23), F = 5.13 (8, 23), p < 0.001. Positivity model: R² = 0.724, adjusted R² = 0.628, residual SE = 2.649 (df = 23), F = 7.54 (8, 23), p < 0.001. Bold p-values indicate statistical significance (p < 0.05). WWTP, wastewater treatment plant; SD, standard deviation.

| Variable | Incidence Model β (SE) | p-value | Positivity Model β (SE) | p-value |
| --- | --- | --- | --- | --- |
| **Population and infrastructure** |  |  |  |  |
| log(Population) | 0.505 (0.134) | **<0.001** | 0.528 (0.116) | **<0.001** |
| Sewered fraction | 0.297 (0.773) | 0.705 | 0.042 (0.488) | 0.933 |
| WWTPs per 100k | 0.066 (0.045) | 0.154 | 0.089 (0.036) | **0.020** |
| Balance index | -0.574 (0.860) | 0.511 | -0.200 (0.679) | 0.771 |
| **Signal variability** |  |  |  |  |
| SD incidence (in-network) | 0.00035 (0.00110) | 0.757 | — | — |
| SD incidence (off-network) | -0.00032 (0.00064) | 0.617 | — | — |
| SD positivity (in-network) | — | — | 8.57 (6.19) | 0.179 |
| SD positivity (off-network) | — | — | -8.98 (3.29) | **0.012** |
| **Surveillance intensity** |  |  |  |  |
| Mean tests (in-network) | -1.49×10<sup>-5</sup> (2.62×10<sup>-5</sup>) | 0.575 | -8.66×10<sup>-6</sup> (2.36×10<sup>-5</sup>) | 0.717 |
| Mean tests (off-network) | $-1.17 \times 10^{-5}$ ($1.16 \times 10^{-5}$) | 0.326 | $-1.73 \times 10^{-5}$ ($9.17 \times 10^{-6}$) | 0.072 |

### Wastewater Concentration as a Sentinel Signal for Off-Network Populations

We correlated weekly SARS-CoV-2 wastewater concentration (log copies/mL) with in-network and off-network clinical indicators within each geographic unit. Paired Wilcoxon signed-rank tests revealed no significant difference between wastewater-to-in-network and wastewater-to-off-network correlations at either scale (county incidence: p = 0.151, N = 15; county positivity: p = 0.169, N = 15; mobility-defined community incidence: p = 0.424, N = 12; mobility-defined community positivity: p = 0.424, N = 12). Deltas were mixed in direction across both frameworks (county median Δ = +0.032 for incidence, −0.030 for positivity; mobility-defined community median Δ = +0.018 for incidence, +0.003 for positivity), with no systematic preference of wastewater signal for the population it directly samples (Figures 4 and 5).

**Figure 4.**
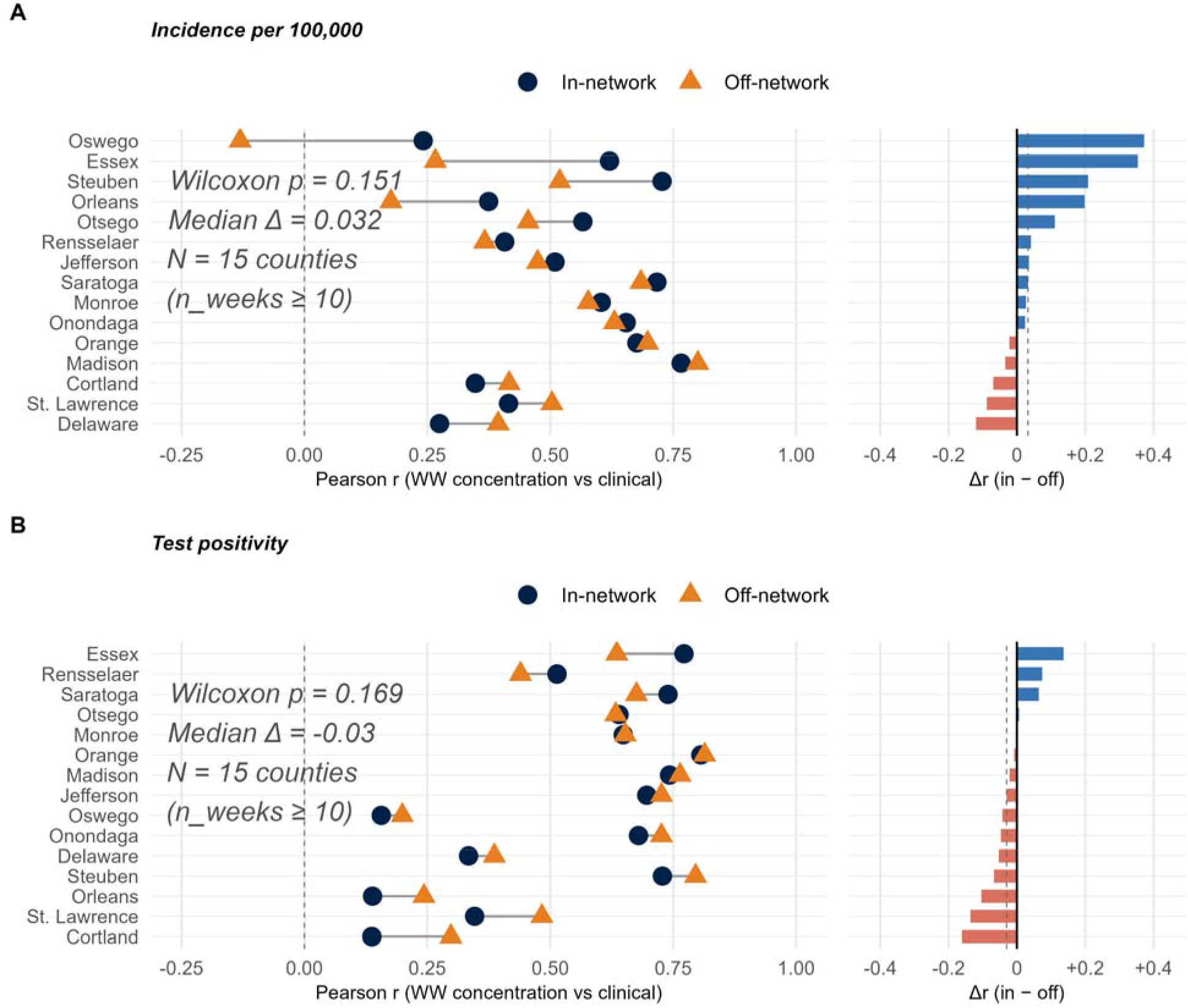
Wastewater correlation with in-network versus off-network clinical burden by county. Pearson correlation coefficients between weekly SARS-CoV-2 wastewater concentration (log copies/mL) and (*A*) incidence per 100,000 and (*B*) test positivity are shown separately for in-network (blue circles) and off-network (orange triangles) populations. Only counties with at least 10 paired weeks of wastewater data are shown (N = 15). The right panel shows Δr (in-network minus off-network); blue bars favor in-network, red bars favor off-network.

**Figure 5.**
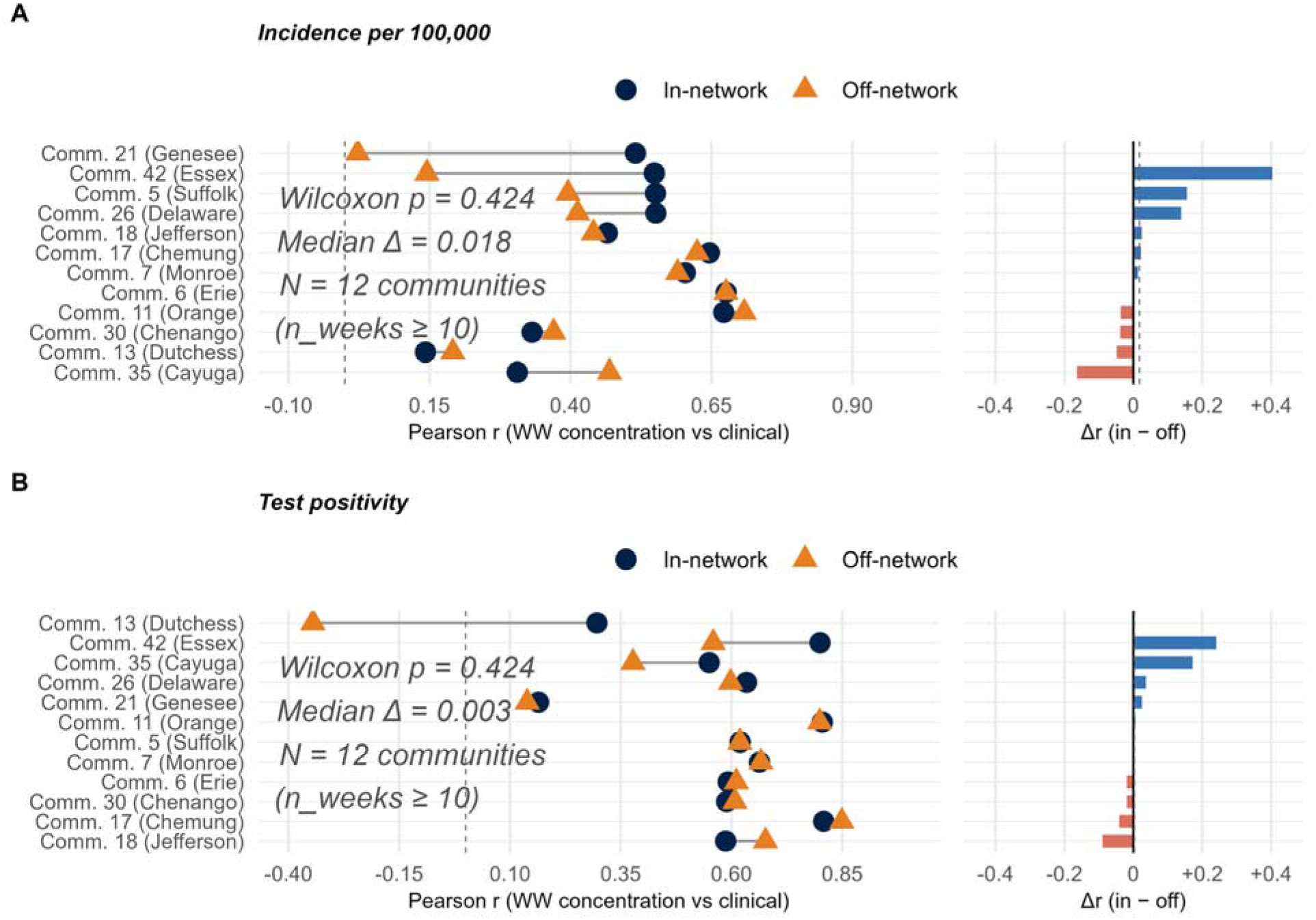
Wastewater correlation with in-network versus off-network clinical burden by mobility-defined community. Pearson correlation coefficients between weekly SARS-CoV-2 wastewater concentration (log copies/mL) and (*A*) incidence per 100,000 and (*B*) test positivity are shown separately for in-network (blue circles) and off-network (orange triangles) populations. Only communities with at least 10 paired weeks of wastewater data are shown (N = 12). The right panel shows Δr (in-network minus off-network); blue bars favor in-network, red bars favor off-network. Mobility-defined communities are labeled by community number and primary county name.

## Discussion

### Representativeness of WWTPs in Magnitude and Timing

These analyses show that WWTPs reflect infectious disease dynamics of mobility-defined communities extending beyond their catchment areas. COVID-19 dynamics were quite similar among on- and off-network populations, supporting the interpretation of WWTPs as representative sentinel sites for surrounding communities. At both the county and mobility-defined community level, in/off-network incidence and test positivity tracked closely in magnitude across all epidemic phases, and cross-correlation analysis confirmed synchronous timing with peak alignment and no lag or lead time (**Figure 1**). This representativeness was further confirmed by the wastewater concentration analysis, which found no significant difference between wastewater-to-in-network and wastewater-to-off-network correlations at either scale or metric (paired Wilcoxon signed-rank tests: county incidence p = 0.151, county-level test positivity p = 0.169, mobility-defined community incidence p = 0.424, mobility-defined community test positivity p = 0.424; Figures 4 and 5). An infectious disease signal from the WWTP is not preferentially informative for the in-network population from which it originates. However, the statewide alignment masked mobility-defined community heterogeneity: smaller, more rural communities showed weaker in/off synchrony, and population size was the dominant structural predictor of alignment at both scales.

### Statewide Coverage Through Representativeness

Because representativeness holds broadly, existing WBE infrastructure provides effective surveillance coverage extending well beyond in-network boundaries. At the county level, where 95% of the counties contained at least some WWTP coverage, median in/off-network incidence correlations were high (r = 0.909), and population size was the dominant structural predictor of alignment. The consistent finding that population size predicts in/off-network synchrony is interpretable through two complementary mechanisms. First, larger communities are likely to have greater spatial overlap and social integration between in-network and off-network residents, who may share workplaces, schools, commercial establishments, and social networks. Under these conditions, transmission dynamics propagate across the in/off-network boundary, producing correlated epidemic curves regardless of differential sewershed connectivity. Second, larger communities generate more stable weekly testing estimates in the in/off-network strata, reducing measurement noise that would otherwise attenuate observed correlations in smaller communities. For operational surveillance purposes, county-level WBE in New York State is sufficient to capture infectious disease dynamics in both connected and off-network populations across most of the state. The corollary is equally important: in smaller counties and mobility-defined communities, off-network populations may experience genuinely divergent dynamics driven by geographic separation between in-network town centers and off-network rural peripheries, differential access to testing and healthcare, distinct social network structures, or localized transmission events in small off-network communities that do not propagate to the broader population. Whatever the mechanism, the implication for wastewater surveillance is significant: in smaller communities, the wastewater signal may be a poor proxy for the epidemic trajectory of the unconnected majority.

The 42% of mobility-defined communities with no WWTP coverage represent only 1.2% of the state population even though WWTP capacity is concentrated in urban centers. Among mixed-coverage communities, the median sewered fraction was only 0.247, meaning fewer than one in four residents were connected to a monitored facility. This pattern reflects longstanding differences in WWTP investment between urban and rural areas, where centralized sewage treatment has historically been prioritized in densely populated areas where per-capita infrastructure costs are lower. Importantly, where WWTPs do exist, they effectively capture disease dynamics in surrounding off-network populations, meaning the surveillance gap is not one of representativeness but of physical infrastructure presence. This gap is particularly concerning given evidence that rural communities experienced distinct COVID-19 trajectories, with rural mortality rates exceeding urban rates during every variant-associated surge (20,21), higher per-capita mortality persisting for up to two years (22), lower vaccination coverage (21), and more limited access to testing infrastructure (23). Expanding wastewater surveillance to smaller treatment facilities and rural communities should be considered a surveillance equity priority, particularly for future infectious disease emergencies where early signal detection in all population subgroups is critical. The natural separation in sewered fraction distribution, with no communities falling in the 0.001–0.05 or 0.95–0.999 boundary zones, suggests that wastewater infrastructure in NYS tends to be deployed at either a mobility-defined community-wide scale or not at all, implying that any increased surveillance investment should prioritize the entirely off-network communities rather than incremental expansion in partially covered areas.

### Counties as Operational Units and the Complementary Value of Mobility-defined communities

County-level WBE performed well in New York State, and for most practical surveillance applications, counties provide adequate analytical units. However, mobility-defined communities offered complementary scientific insight. Although county-level correlations were somewhat higher in aggregate (median r = 0.909 versus 0.877), the mobility-defined community framework offered better population balance between in/off-network strata (87.5% versus 69.8% in balanced range) and a higher floor on representativeness (minimum r = 0.440 versus 0.177). Importantly, the mobility-defined community framework revealed infrastructure predictors, specifically WWTP density and off-network signal variability, that were not detectable at the county level, suggesting that wastewater infrastructure characteristics are actionable determinants of representativeness visible only when geographic units align with population movement patterns. County boundaries, drawn for governance rather than epidemiological purposes, typically do not align with functional population boundaries and may combine communities that are not connected through routine movement (24). This extends prior work demonstrating that mobility-informed community structures more accurately capture population connectivity relevant to infectious disease transmission than static administrative units (25). Mobility-based frameworks address this mismatch but require access to mobile device location data, which may not be available in all jurisdictions or surveillance contexts. As wastewater surveillance expands beyond COVID-19 to other respiratory diseases and antimicrobial resistance monitoring, grounding system design in mobility-defined catchment populations could improve the equity and accuracy of population-level disease estimates, while county frameworks remain the practical default where mobility data are unavailable.

This study has several limitations. The subtraction approach assumes similar testing propensity between in-network and off-network residents, which may bias incidence correlations upward in areas with differential testing access. However, test positivity inherently controls for this bias, and the consistently strong positivity correlations suggest testing access alone does not account for the observed alignment. Sewershed population denominators were derived from plants with capacity ≥ 1 MGD, which by design excludes approximately 400 smaller facilities; however, the included approximately 200 plants serve an estimated 95% of New York State’s total in-network population, so the misclassified population is small relative to the total off-network denominator. The pooled analysis spans the Alpha, Delta, and early Omicron variant waves, and future work should examine whether these patterns hold across distinct epidemic phases. These findings may generalize to other respiratory and enteric pathogens sharing movement-mediated transmission but may not hold for vector-borne or sexually transmitted infections where transmission depends on vector habitat or sexual network structure rather than routine mobility. Finally, this analysis is specific to New York State outside NYC, and replication in other geographic contexts with different settlement geographies and sewershed coverage patterns would strengthen generalizability.

Further consideration concerns the temporal dynamics of WBE signal detection relative to clinical case ascertainment. Wastewater surveillance detects SARS-CoV-2 RNA shed by infected individuals prior to symptom onset and clinical testing, providing potential lead time over clinical surveillance. However, this lead time advantage applies directly only to the in-network population. While wastewater signals may indirectly reflect off-network dynamics through routine human movement, off-network populations have no dedicated, early warning mechanism of their own. These delays were epidemiologically meaningful, with cumulative prevalence increasing five- to eleven-fold within a week after detection under higher transmission or reduced assay sensitivity (17). In communities where off-network populations comprise the majority, as in 42% of NYS mobility-defined communities in this study, the absence of an equivalent early warning mechanism for off-network residents compounds the surveillance equity gap identified here and warrants explicit consideration in WBE surveillance design (26).

### Conclusions

WWTPs broadly represent infectious disease dynamics in surrounding off-network populations in both magnitude and timing, supporting their use as sentinel surveillance sites for in-network and off-network communities alike. Because this representativeness holds across most counties, existing WBE infrastructure effectively covers the vast majority of New York State’s population, including those not directly connected to the sewer network. County-level WBE frameworks are operationally sufficient, while mobility-defined communities offer complementary analytical value for identifying where and why representativeness varies. As WBE matures beyond COVID-19 into routine multi-pathogen surveillance, incorporating both perspectives into system design is a tractable step toward more equitable and epidemiologically valid infectious disease surveillance.

## Methods

We evaluated whether WWTPs represent infectious disease dynamics in off-network communities using two complementary analytical frameworks applied to COVID-19 clinical testing data from New York State (NYS) (January 2021–April 2022). First, we conducted the analysis at the county level as a baseline. Second, we applied a community detection algorithm to census tract-level human mobility networks to define mobility communities that reflect functional population connectivity rather than administrative boundaries and repeated the analysis within these mobility-defined units. New York City’s five boroughs (Bronx, Kings, New York, Queens, and Richmond counties) were excluded from all analyses due to their distinct in-network infrastructure, population density, and public health surveillance systems.

### Mobility data

We characterized human movement between in-network and off-network communities using anonymized, aggregated mobile device location records from Advan Research (now provided by Dewey, San Francisco, CA) (27). These data record weekly visits from inferred home census block groups (CBGs) to points of interest (POIs) and have been widely used to quantify transmission-relevant population mixing and spatial heterogeneity in infectious disease dynamics (28,29). Weekly records were restricted to NYS and aggregated to the month level. We expanded visitor home CBG distributions into origin–destination flows, and both origins and destinations were converted to census tracts by truncating 12-digit GEOIDs to 11 digits. We also obtained monthly tract–tract flows by summing visits across all weeks. In the analysis we included only NYS resident origins, and tract pairs with fewer than five visits per month were excluded to reduce noise. The census tract populations were obtained from the 2020 ACS (variable B01003_001) and linked to mobility communities via tract-level GEOID using the tidycensus package (v1.7.1)(30).

Two mobility-defined communities corresponding to the Allegany Territory of the Seneca Nation (Cattaraugus County) were excluded from all analyses prior to modeling. The first mobility-defined community (census tract 9402) recorded zero population in the 2020 ACS, had no associated COVID-19 clinical testing records, and contained no sewershed coverage, consistent with its status as an unpopulated boundary designation within tribal lands. The second mobility-defined community (census tract 9400) recorded 201 residents but exhibited a disproportionately sparse testing pattern inconsistent with New York State’s clinical testing infrastructure used by all other communities in the dataset (mean 9.71 tests per week; 41 of 68 weeks with fewer than 10 tests), reflecting the Seneca Nation’s independent COVID-19 testing program rather than the state-administered surveillance system. Both tracts are enumerated under separate tribal census and health protocols and are not directly comparable to the general population of communities comprising the remainder of the analytical sample. Because each of these tracts constituted its own mobility-defined community under the Walktrap algorithm, their exclusion reduced the analytical sample from 66 to 64 mobility-defined communities.

### Community detection algorithm

Community detection methods identify clusters of densely connected nodes within a network, with algorithm choice materially affecting results through differences in computational cost, resolution, and optimization criteria (31). To select an appropriate algorithm for our mobility network, we characterized the network following the empirical benchmarking framework of Yang et al. (2016) (31), who evaluated eight state-of-the-art community detection algorithms across a wide range of network sizes (N) and mixing parameters (μ) using the Lancichinetti-Fortunato-Radicchi (LFR) benchmark. The mixing parameter μ, which is defined as the fraction of edges crossing mobility-defined community boundaries, is the most influential determinant of algorithm performance (31). Our mobility-defined community network comprised N = 64 populated communities (nodes) with 226 geographic adjacency edges (mean degree = 6.95). The mixing parameter was estimated using the two-step procedure recommended by Spinglass (31) and multilevel algorithms were applied as preliminary estimators, yielding μ = 0.447 and μ = 0.465 respectively, confirming a robust estimate of μ ≈ 0.45. With N < 1,000 and μ < 0.5, our network falls within the region where multiple algorithms perform reliably (31).

Walktrap was selected over alternatives for three reasons. First, its accuracy turning point extends past μ = 0.5, providing a margin of robustness given that our estimated μ ≈ 0.45 lies close to this threshold, a property not shared by Label propagation or Infomap, whose accuracy drops sharply at or before μ = 0.5. Second, unlike Label propagation, Walktrap produces stable community assignments with low variance across repeated runs, an important property for a geographically fixed network analyzed across weekly time steps. Third, the Multilevel algorithm’s modularity-based optimization is subject to a known resolution limit (25) that systematically merges small communities into larger ones. This was directly observed in our preliminary estimation, where Multilevel collapsed 65 communities into 6 macro-regions, obscuring precisely the small rural communities central to our primary research question.

We identified mobility-defined communities by applying the Walktrap algorithm (32) implemented in the igraph R software package (33) to a network of 2,870 New York State census tracts outside NYC, yielding 66 mobility-defined communities (64 after exclusion of two Seneca Nation tracts), as represented in Eq. 1.

|  |  |
| --- | --- |
| $P_{ij} = \frac{W_{ij}}{\sum_k W_{ik}}$ | [Eq.1]. In our weighted tract mobility network $G = (V, E)$, each node $V$ represents a census tract and each edge weight $W_{ij}$ corresponds to the total visits between tracts $i$ and $j$, with edge weights treated as transition intensities for a 4-step random walk. $P_{ij}$ represents the probability of moving from tract $i$ to tract $j$, where the denominator sums the edge weights from tract $i$ to all adjacent tracts $k$. |

### Delineation of in-network and off-network populations

To align wastewater infrastructure with mobility-defined populations, we dissolved census tracts belonging to the same mobility-defined community to form community polygons. The WWTPs sewershed boundaries were spatially overlaid with these polygons, and each sewershed was assigned to the mobility-defined community containing the largest proportion of its service area. The in-network population for each mobility-defined community was estimated by summing the reported population served by each included WWTP assigned to that mobility-defined community; these figures are self-reported by treatment plant operators and do not rely on spatial interpolation or assumptions about uniform population distribution within census tracts. Total mobility-defined community population was calculated by summing 2020 ACS tract-level estimates (variable B01003_001) for all tracts assigned to each mobility-defined community. The off-network population was defined as the difference between total mobility-defined community population and in-network population.

### Clinical data

We obtained daily COVID-19 clinical testing data from the New York State Department of Health including total tests administered and positive cases per county per day from January 1, 2021 to April 30, 2022 geocoded along two parallel geographic frameworks. In the county framework, each test was classified as in-network (residential address within the catchment area of a WWTP permitted to discharge at least 1.0 million gallons of wastewater per day) (34), or off-network (within the county but outside any included WWTP catchment). In the mobility-defined community framework, the same in-network/off-network classification was applied using the sewershed-to-community spatial assignments described above. We aggregated COVID- 19 tests conducted and COVID-19 positive tests of these conditions to weekly totals aligned to epidemiological weeks. In-network clinical counts were obtained by summing sewershed-level testing data within each county or mobility-defined community, and off-network counts were calculated by subtracting in-network totals from overall county or mobility-defined community clinical testing totals. The incidence per 100,000 population and test positivity were calculated separately for in-network and off-network populations using their respective population denominators.

### County-level analysis

At the county level, in-network population denominators were defined as the total population served by included WWTPs (≥1 MGD capacity) within each county, derived from sewershed population statistics. Off-network population was calculated as the difference between the county’s total population (American Community Survey 2020 5-year estimates) and the in-network population. Clinical testing data were filtered to include only New York State counties, excluding records from neighboring states that appeared in the geocoded dataset due to cross-border testing. Counties were classified as mixed-coverage (sewered fraction >0.001 and <0.999), fully in-network (≥0.999), or entirely off-network (≤0.001). Weekly in-network and off-network incidence per 100,000 and test positivity were computed for each county, and Pearson correlations between in/off-network time series were calculated for each mixed-coverage county using weekly aggregated data (minimum three paired weeks; test positivity gated at ≥50 tests per stratum); two counties with zero-variance or near-zero in-network series were excluded, yielding N = 53 for incidence and N = 52 for positivity correlations. Statewide correlations were computed by aggregating all county-level counts weekly and recalculating rates using summed population denominators. Cross-correlation functions were examined at lags of ±6 weeks to assess temporal alignment.

### Mobility-defined community analysis

The mobility-defined community analysis followed the same analytical framework as the county level, with in-network and off-network populations defined using the sewershed-to-community spatial assignments described above. Weekly in-network clinical counts were aggregated from sewershed-level testing data, and off-network counts were derived by subtracting in-network totals from mobility-defined community clinical totals. Because sewershed catchment boundaries and mobility community boundaries are independently defined, minor spatial misalignment between the two occasionally produced community-weeks where geocoded in-network counts slightly exceeded community totals; in these instances, in-network counts were constrained to community totals to maintain non-negative off-network estimates.

Communities were classified using the same coverage thresholds as the county analysis. Pearson correlations were computed for each mixed-coverage community (minimum three paired weeks; test positivity gated at ≥50 tests per stratum), with statewide aggregations restricted to the 32 mixed-coverage communities. Wastewater concentration analyses were restricted to counties and communities with a minimum of 10 paired weeks of concentration and clinical data (county N = 15 of 53 mixed-coverage counties; mobility-defined community N = 12 of 32 mixed communities), reflecting sites with sufficient surveillance continuity for stable correlation estimation. Analyses were restricted to mixed-coverage units where both in-network and off-network populations could be independently estimated.

### Factors affecting correlation between clinical indicators of COVID-19 transmission among on- and off-network communities

To identify mobility-defined community structural characteristics associated with the strength of epidemiological alignment between in/off-network populations, we fit weighted linear regression models separately for incidence per 100,000 population and test positivity. Analyses were restricted to the 32 mixed-coverage communities (sewered fraction > 0.001 and < 0.999), where both in/off-network COVID-19 burden could be independently estimated by subtraction. Community type clustering was robust to alternative threshold definitions with no communities falling within the 0.001–0.05 /0.95–0.999 boundary zones, confirming natural separation between community types in the sewered fraction distribution. The dependent variable in each model was the Fisher z-transformed Pearson correlation coefficient between weekly in/off-network COVID-19 metrics, computed over the full study period for each mobility-defined community. The Fisher transformation was applied to stabilize the variance of correlation coefficients and enable valid inference across communities with differing numbers of observation weeks using Eq. 2.

|  |  |
| --- | --- |
| $Z_r = 1/2 \ln \left[ \frac{1+r}{1-r} \right]$ | [Eq.2]. Fisher z-transformation of the Pearson correlation coefficient $r$, where $Z_r$ is the transformed outcome used as the dependent variable in weighted linear regression models. Models were weighted by $w_i = \max(1, n_i)$, where $n_i$ is the number of completely paired observation weeks per mobility-defined community or county. |

Models were weighted by the number of completely paired mobility-defined community observation weeks ( w*_i=_*max (1,n*i*). This weighting scheme down-weighted communities with sparse time series, reducing the influence of unstable correlation estimates on model inference. Eight candidate predictors were included in the full models: log-transformed total mobility-defined community population, sewered fraction, WWTPs per 100,000 population, balance index (defined as min (sewered fraction, 1 − sewered fraction), capturing the degree of mixing between in-network and off-network subpopulations), standard deviation and mean tests of weekly in/off burden. A parsimonious positivity model retaining only statistically significant predictors from the full positivity model was additionally fitted as a sensitivity analysis. Model fit was assessed using, adjusted, residual standard error, and overall *F*-statistic. Analyses were conducted in the statistical package R, version 4.4.1 (35).

## Data Availability

COVID-19 clinical testing data were obtained from the New York State Department of Health through a data sharing agreement and are not publicly available. Wastewater concentration data are available through the New York State Wastewater Surveillance Network. Mobility data were obtained from Advan Research (now provided by Dewey, San Francisco, CA) and are available to academic researchers upon request at https://www.deweydata.io. Analysis code will be made available upon acceptance.

## Acknowledgments

This work was supported by the Centers for Disease Control and Prevention (CDC) Epidemiology and Laboratory Capacity for Prevention and Control of Emerging Infectious Diseases (ELC) program, New York State unique federal award number NU50CK000516 (PI: David A. Larsen). Clinical COVID-19 testing data were provided by the New York State Department of Health. Mobility data were provided by Advan Research through the Dewey Academic Data Program. Wastewater surveillance data were provided through the New York State connected to WWTPs with permitted capacity ≥1 MGD. The content is solely the responsibility of the authors and does not necessarily represent the official views of the funding agencies.

## References

[1] T. Pöyry, M. Stenvik, and T. Hovi, “Viruses in sewage waters during and after a poliomyelitis outbreak and subsequent nationwide oral poliovirus vaccination campaign in Finland,” Appl. Environ. Microbiol., vol. 54, no. 2, pp. 371–374, Feb. 1988, doi: 10.1128/aem.54.2.371-374.1988.

[2] W. Ahmed et al., “Decay of SARS-CoV-2 and surrogate murine hepatitis virus RNA in untreated wastewater to inform application in wastewater-based epidemiology,” Environ. Res., vol. 191, p. 110092, Dec. 2020, doi: 10.1016/j.envres.2020.110092.

[3] A. Bivins et al., “Wastewater-Based Epidemiology: Global Collaborative to Maximize Contributions in the Fight Against COVID-19,” Environ. Sci. Technol., vol. 54, no. 13, pp. 7754–7757, Jul. 2020, doi: 10.1021/acs.est.0c02388.

[4] B. Hughes et al., “Respiratory Syncytial Virus (RSV) RNA in Wastewater Settled Solids Reflects RSV Clinical Positivity Rates,” Environ. Sci. Technol. Lett., vol. 9, no. 2, pp. 173– 178, Feb. 2022, doi: 10.1021/acs.estlett.1c00963.

[5] M. K. Wolfe et al., “Wastewater-Based Detection of Two Influenza Outbreaks,” Environ. Sci. Technol. Lett., vol. 9, no. 8, pp. 687–692, Aug. 2022, doi: 10.1021/acs.estlett.2c00350.

[6] P. Kilaru et al., “Wastewater Surveillance for Infectious Disease: A Systematic Review,” *Am. J. Epidemiol.*, p. kwac175, Oct. 2022, doi: 10.1093/aje/kwac175.

[7] D. Gerrity, K. Papp, M. Stoker, A. Sims, and W. Frehner, “Early-pandemic wastewater surveillance of SARS-CoV-2 in Southern Nevada: Methodology, occurrence, and incidence/prevalence considerations,” Water Res. X, vol. 10, p. 100086, Jan. 2021, doi: 10.1016/j.wroa.2020.100086.

[8] J. Weidhaas et al., “Correlation of SARS-CoV-2 RNA in wastewater with COVID-19 disease burden in sewersheds,” Sci. Total Environ., vol. 775, p. 145790, Jun. 2021, doi: 10.1016/j.scitotenv.2021.145790.

[9] R. A. Chowdhury et al., “Effectiveness of Wastewater-Based Epidemiology as an Early Warning Tool to Detect SARS-CoV-2 (COVID-19),” Health (N. Y.), vol. 16, no. 07, pp. 635–656, 2024, doi: 10.4236/health.2024.167045.

[10] T. Prado et al., “Wastewater-based epidemiology as a useful tool to track SARS-CoV-2 and support public health policies at municipal level in Brazil,” Water Res., vol. 191, p. 116810, Mar. 2021, doi: 10.1016/j.watres.2021.116810.

[11] Q. Yu, S. W. Olesen, C. Duvallet, and Y. H. Grad, “Assessment of sewer connectivity in the United States and its implications for equity in wastewater-based epidemiology,” *PLOS Glob*. Public Health, vol. 4, no. 4, p. e0003039, Apr. 2024, doi: 10.1371/journal.pgph.0003039.

[12] S. A. Hall, J. S. Kaufman, and T. C. Ricketts, “Defining Urban and Rural Areas in U.S. Epidemiologic Studies,” J. Urban Health, vol. 83, no. 2, pp. 162–175, Apr. 2006, doi: 10.1007/s11524-005-9016-3.

[13] D. Khanna et al., “Rural-urban disparity in cancer burden and care: findings from an Indian cancer registry,” BMC Cancer, vol. 24, no. 1, p. 308, Mar. 2024, doi: 10.1186/s12885-024-12041-y.

[14] G. O’Reilly, D. O’ Reilly, M. Rosato, and S. Connolly, “Urban and rural variations in morbidity and mortality in Northern Ireland,” BMC Public Health, vol. 7, no. 1, p. 123, Dec. 2007, doi: 10.1186/1471-2458-7-123.

[15] N. J. MacKinnon et al., “Mapping Health Disparities in 11 High-Income Nations,” *JAMA Netw*. Open, vol. 6, no. 7, p. e2322310, Jul. 2023, doi: 10.1001/jamanetworkopen.2023.22310.

[16] L. Xiang, J. W. Keck, J. Gallimore, A. Sakhaei, E. Loh, and S. M. Berry, “Wastewater Infrastructure as a Public Health Tool: Agent-Based Modeling of Surveillance Strategies in a COVID-19 Context,” Systems, vol. 13, no. 12, p. 1093, Dec. 2025, doi: 10.3390/systems13121093.

[17] N. DelaPaz-Ruíz, E.-W. Augustijn, M. Farnaghi, S. A. Abdulkareem, and R. Zurita Milla, “Integrating agent-based disease, mobility and wastewater models for the study of the spread of communicable diseases,” Geospatial Health, vol. 20, no. 1, Feb. 2025, doi: 10.4081/gh.2025.1326.

[18] H. Amiri, A. Deverakonda, Y. Wang, and A. Züfle, “Where do We Poop? City-Wide Simulation of Defecation Behavior for Wastewater-Based Epidemiology,” Jan. 03, 2026, arXiv: arXiv:2601.04231. doi: 10.48550/arXiv.2601.04231.

[19] X. Hou et al., “Intracounty modeling of COVID-19 infection with human mobility: Assessing spatial heterogeneity with business traffic, age, and race,” Proc. Natl. Acad. Sci., vol. 118, no. 24, p. e2020524118, Jun. 2021, doi: 10.1073/pnas.2020524118.

[20] R. Saelee et al., “Disparities in COVID-19 Vaccination Coverage Between Urban and Rural Counties — United States, December 14, 2020–January 31, 2022,” MMWR Morb. Mortal. Wkly. Rep., vol. 71, no. 9, pp. 335–340, Mar. 2022, doi: 10.15585/mmwr.mm7109a2.

[21] S. H. Cross, R. M. Califf, and H. J. Warraich, “Rural-Urban Disparity in Mortality in the US From 1999 to 2019,” JAMA, vol. 325, no. 22, p. 2312, Jun. 2021, doi: 10.1001/jama.2021.5334.

[22] J. T. Mueller, K. McConnell, P. B. Burow, K. Pofahl, A. A. Merdjanoff, and J. Farrell, “Impacts of the COVID-19 pandemic on rural America,” Proc. Natl. Acad. Sci., vol. 118, no. 1, p. 2019378118, Jan. 2021, doi: 10.1073/pnas.2019378118.

[23] Y. Zhu, D. T. Hill, Y. Zhou, and D. A. Larsen, “The effect of the modifiable areal unit problem (MAUP) on spatial aggregation of COVID-19 wastewater surveillance data,” Sci. Total Environ., vol. 957, p. 177676, Dec. 2024, doi: 10.1016/j.scitotenv.2024.177676.

[24] S. Fortunato and M. Barthélemy, “Resolution limit in community detection,” Proc. Natl. Acad. Sci., vol. 104, no. 1, pp. 36–41, Jan. 2007, doi: 10.1073/pnas.0605965104.

[25] M. Neyra Blatz et al., “Equities and Inequities Inherent in Wastewater Surveillance Systems for Public Health: New York State, 2020–2024,” Am. J. Public Health, pp. e1–e9, May 2026, doi: 10.2105/AJPH.2026.308472.

[26] Advan Research, “Foot Traffic / Weekly Patterns Plus.” Dewey Data. doi: 10.82551/C103-N851.

[27] S. Chang et al., “Mobility network models of COVID-19 explain inequities and inform reopening,” Nature, vol. 589, no. 7840, pp. 82–87, Jan. 2021, doi: 10.1038/s41586-020-2923-3.

[28] N. Oliver et al., “Mobile phone data for informing public health actions across the COVID- 19 pandemic life cycle,” Sci. Adv., vol. 6, no. 23, p. eabc0764, Jun. 2020, doi: 10.1126/sciadv.abc0764.

[29] Walker K, Herman M. tidycensus, ALoad US Census Boundary and Attribute Data as “tidyverse” and ‘sf’ -Ready Data Frames., R package version 1.7.1. [Online]. Available: https://walker-data.com/tidycensus/

[30] Z. Yang, R. Algesheimer, and C. J. Tessone, “A Comparative Analysis of Community Detection Algorithms on Artificial Networks,” Sci. Rep., vol. 6, no. 1, p. 30750, Aug. 2016, doi: 10.1038/srep30750.

[31] P. Pons and M. Latapy, “Computing communities in large networks using random walks (long version),” Dec. 12, 2005, arXiv: arXiv:physics/0512106. doi: 10.48550/arXiv.physics/0512106.

[32] Csardi, G. & Nepusz, T.,

[33] D. T. Hill and D. A. Larsen, “Using geographic information systems to link population estimates to wastewater surveillance data in New York State, USA,” *PLOS Glob*. Public Health, vol. 3, no. 1, p. e0001062, Jan. 2023, doi: 10.1371/journal.pgph.0001062.

[34] R Core Development Team, “R: A Language and Environment for Statistical Computing,” HttpwwwR-Proj., 2010.

